# IVA, a locally deployable Intelligent Variant Analyser based on Small Language Model for complete case-level and trio-based genomic interpretation

**DOI:** 10.64898/2026.10.07.26364945

**Authors:** Eric Ducret, Jing Zhai, Sergey Oreshkov, Abdur Rashid, Helen Frederiksen, Mukhtar Ullah, Alexandre Laverre, Romain Forey, Cedric Le Caignec, Muhammad Ansar, Federico Santoni

## Abstract

**Background:** Recent work has brought large language models (LLMs) into clinical variant interpretation. In most existing approaches, the model is confined to text-level operations, retrieving, extracting, compacting, and summarizing heterogeneous evidence, while the genuine genetic evaluation is delegated to a separate deterministic engine. Other tools employ LLMs to mine functional evidence for a single ACMG criterion, to rerank a classifier’s output, or to answer isolated variant questions. Integrative genetic reasoning, by contrast, demands multi-source evidence weighing for each variant, reconciliation of inheritance with observed zygosity and parental genotypes, joint interpretation of variants in the same gene, and the derivation of a case-level conclusion. Moreover, to protect patient privacy, this must be done on hospital premises.

**Methods:** o build an end-to-end variant interpreter that runs on local infrastructure, we benchmarked 13 small language models (SLMs) on 150 multiple-choice genetics questions and selected Qwen3.5-9B. In IVA (Intelligent Variant Analyser), a four-stage pipeline, the model assigns ACMG/AMP criteria, applies de novo, dominant, recessive, and X-linked inheritance logic to trios, and writes a structured report, while deterministic code handles retrieval, phasing, point summation, citation checking, and formatting. We evaluated IVA on synthetic scenarios, 16 solved trios, and 327 solved singleton cases of visual impairment, on a single GPU.

**Results:** On the benchmark, Qwen3.5-9B reached a mean accuracy of 0.81, compared with 0.89 for a 1.6-trillion-parameter cloud model. In the 16 trios, IVA reported every previously identified pathogenic, likely pathogenic, or uncertain-significance variant of interest (sensitivity 100%; precision 39.5%). In the 327 singleton cases, it recovered 93% of the expert-reported causative variants and classified 82% of them as pathogenic or likely pathogenic (precision 43%). Failures fell into four recurrent classes: omission, unverified mechanism, mismatch between reasoning and label, and arithmetic errors.

**Conclusions:** A small language model running on local hardware can serve as the reasoning layer for case-level and trio-based variant interpretation when it is surrounded by deterministic safeguards. Because its outputs are not deterministic, analyst review and prospective multicenter validation remain necessary.

**Description:** IVA is a locally deployable pipeline in which a sub-10B small language model assigns ACMG/AMP criteria, reasons over free text phenotype descriptions, familial inheritance, and writes the clinical report for a complete candidate-variant table, with all model inference running on hospital hardware.

## Introduction

Since next-generation sequencing became routine, the main bottleneck of clinical genetics has shifted from data generation to data interpretation. Even with dedicated software, analysts must query independent databases, apply standardized classification criteria, and synthesize heterogeneous evidence into a conclusion for each patient, a process that is time-consuming and difficult to scale^1^. The ACMG/AMP guidelines^2^ and their refinements^3^ provide the framework, but rule-based pipelines struggle with unstructured evidence^1,4^ as shown by nine laboratories applying the same guidelines to the same variants reached only 34% concordance^5^.

Large language models (LLMs) extract knowledge from narrative text and, within agentic architectures, improve performance across clinical tasks^6^, but general-purpose models often underperform in clinical genetics^7^. Cloud-hosted models also expose protected health information to privacy risks^8^ and reduce institutional control over behavior, versioning, and traceability^9^. Conversely, small language models (SLMs) run on standard institutional hardware, keep patient data in a controlled environment, and make the many sequential calls of agentic workflows affordable^10^, although tool selection and structured reasoning degrade below a critical model size^11^.

Recent systems apply language models to complementary tasks. GeneGPT augments question answering with NCBI tools^12^; AcmGENTIC and AutoPM3 extract evidence for single ACMG criteria from the literature^13,14^; AI-CURA assigns criteria with deterministic tools and uses an LLM only to summarize literature for the criteria that need it^15^; DeepRare and G.AI rank candidate diseases or variants without assigning ACMG criteria^16,17^; DiagAI shortlists variants with a conventional machine-learning model^18^; and MARRVEL-MCP gives LLMs structured access to genomic databases, showing that mid-sized models with tool access approach much larger ones on expert-curated questions^19^.

Two gaps remain. First, existing systems either leave classification to deterministic logic or handle one criterion or one variant at a time; none, to our knowledge, uses a small model on local hospital infrastructure as the integrative reasoning unit over a complete candidate-variant table. Second, joint interpretation of variants within a gene, reconciliation of inheritance with zygosity and parental genotypes, and generation of a complete ACMG-aligned report remain unaddressed, although singletons and trios make up much of the clinical workload. We describe IVA, a pipeline built on this premise, evaluate it on synthetic and real diagnostic cases, and document the safeguards that make a small model usable for this task.

## Methods

### Model selection

We wrote 150 four-option questions, drafted with LLM assistance under domain-expert supervision and then revised, labeled with seven competences: ACMG classification, epigenetics and regulation, genomic diagnostics, inheritance mechanisms, protein function, splicing and RNA, and variant mechanism. Each question was run under all 24 answer orderings (3,600 calls per model), which separates knowledge from sensitivity to option position^20^. We computed mean accuracy; strict accuracy (correct under all 24 orderings); confidence consistency, the stability of option probabilities across orderings, based on the Permutation Bias Metric^21^; choice consistency, the same construct on discrete answers, which also applies to cloud models without token probabilities; and the inverted Brier score^22^. We evaluated 13 instruction-tuned models of 0.8 to 9 billion parameters from six families, including three clinical Phi-3 variants, with DeepSeek-V4-Pro as a cloud reference, using HuggingFace Transformers on one A100 (40 GB) with extended reasoning disabled.

### IVA pipeline

VA is a hybrid framework in which a small language model with greed deconding (temperature 0) performs the interpretive reasoning while deterministic code retrieves the evidence, computes segregation, enforces the ACMG/AMP rules, and checks every model output. IVA accepts an annotated variant table (CSV, TSV, or Excel) and a free-text phenotype; the model maps heterogeneous column conventions to a canonical schema. The analysis proceeds in four stages (Figure 1; Supplementary Methods).

**Figure 1.**
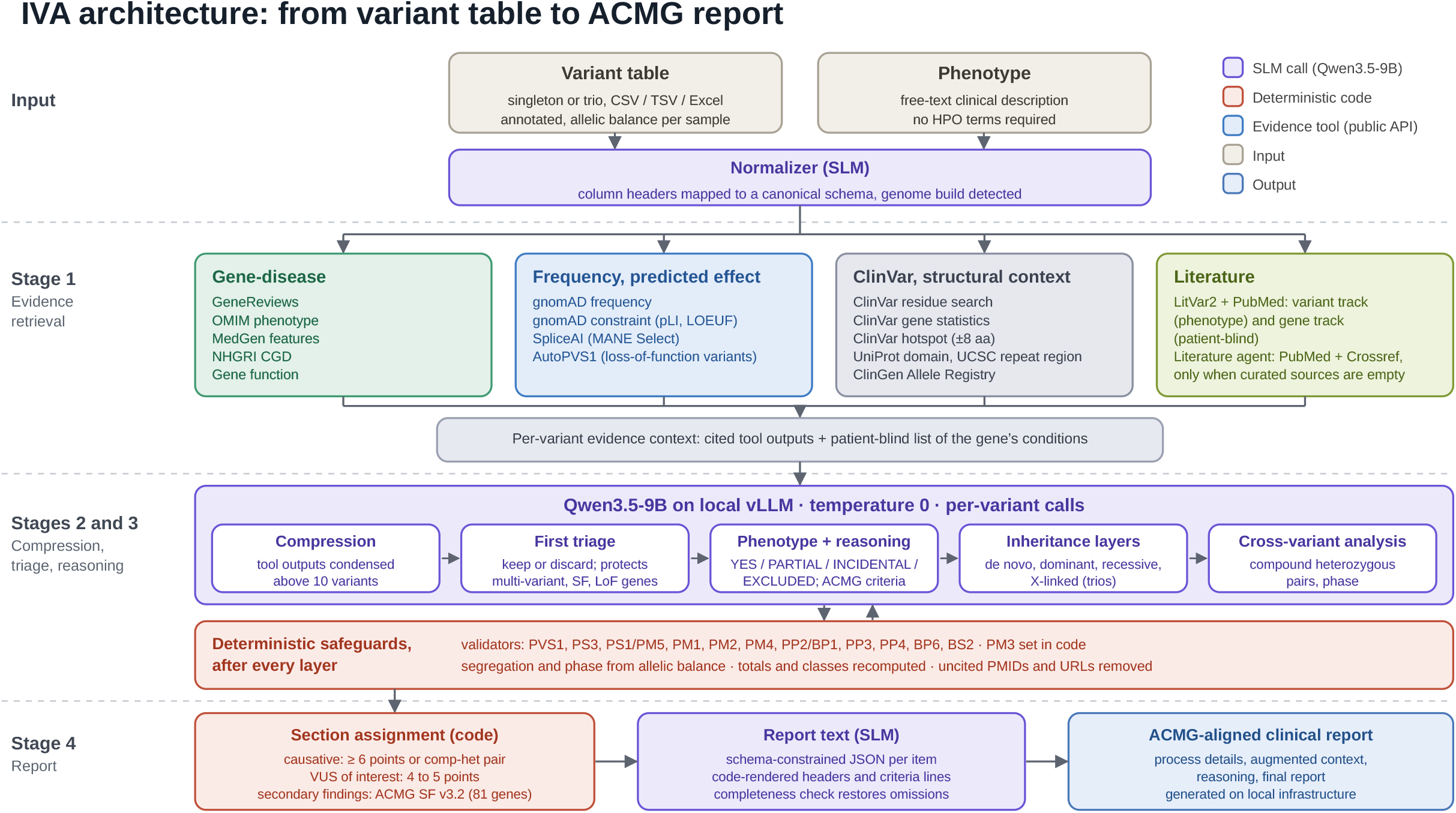
Architecture of IVA. Stage 1 retrieves evidence for each variant from public resources. In Stages 2 and 3, Qwen3.5-9B, running locally, triages the variants, assigns a phenotype verdict and ACMG/AMP criteria, and applies inheritance models in trios, while code recomputes points and validates criteria after every layer (Table S1). In Stage 4, code assigns each variant to a report section and the model writes the text. Purple denotes model calls; red, deterministic code.

#### Stage 1 (retrieval)

For each variant, tools run in a fixed order, from fully deterministic to model-driven: SpliceAI^23^ predictions on the MANE Select transcript, population frequency and gene constraint from gnomAD^24^, residue- and gene-level statistics from ClinVar^25^, ClinGen allele registration^26^, GeneReviews^27^, UniProt^28^ and MedGen^29^ annotations, and, for loss-of-function variants, AutoPVS1^30^. Literature is retrieved from LitVar2^31^ and PubMed through a variant-level track tied to the phenotype and a patient-blind gene-level track based on the gene’s OMIM or CGD^32^ conditions. When no curated source describes a gene, a literature agent searches PubMed and, through Crossref, preprint servers (Table S1; Figure S1). Finally, a separate model call, blinded to the patient, lists every condition that the evidence attributes to the gene, so that the patient’s phenotype cannot leak into this list (Figure 2).

**Figure 2.**
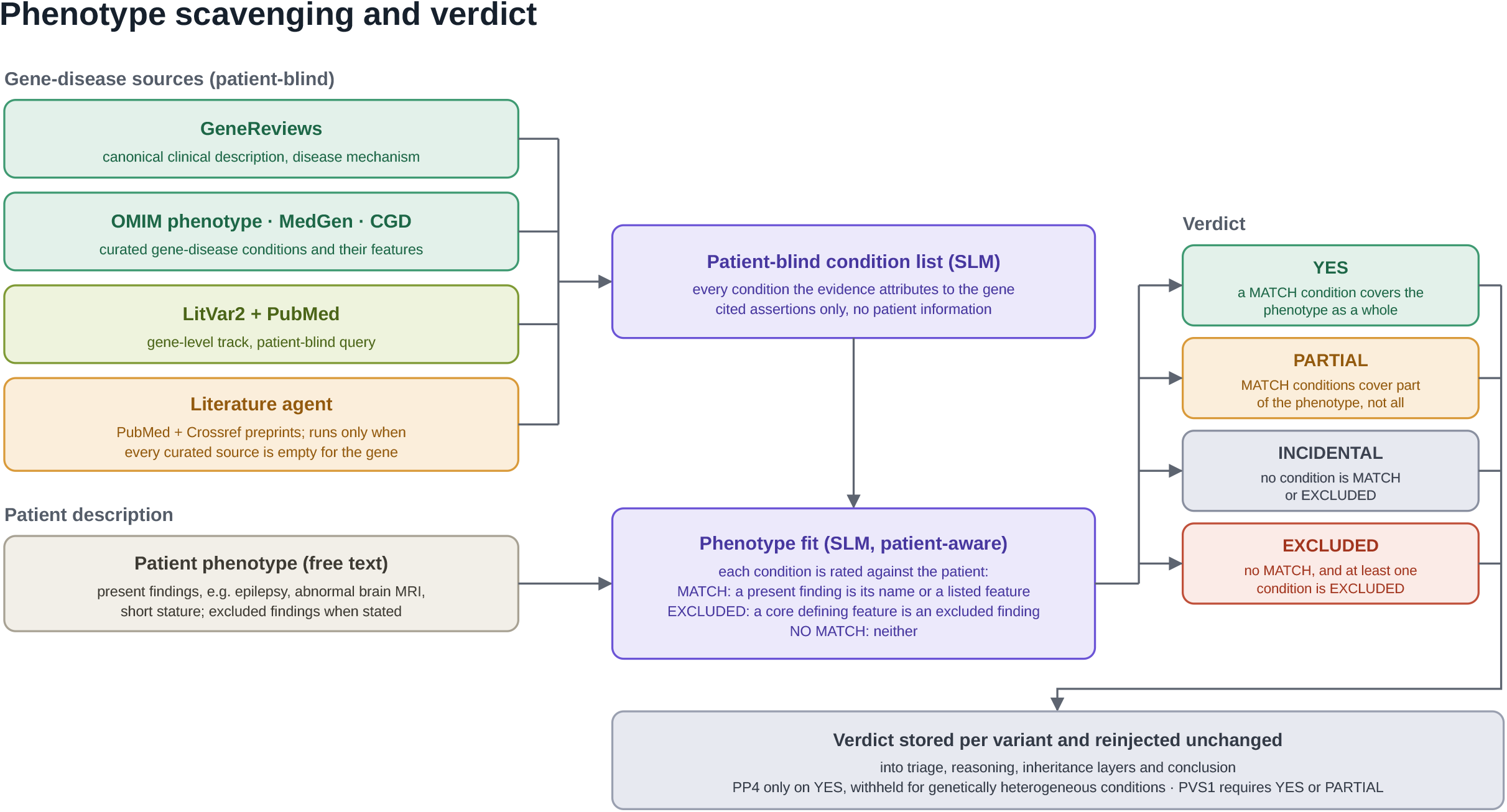
Phenotype Scavenging and Verdict. A patient-blind model call lists the conditions attributed to the gene, and a patient-aware call compares them with the phenotype to return one of four verdicts. The verdict is passed unchanged to later stages and gates PP4 (YES only) and PVS1 (YES or PARTIAL).

#### Stages 2 and 3 (compression, triage, reasoning)

To fit large cases into the context window, the model condenses each tool output once a case exceeds ten variants (twenty for four tools), with a deterministic truncation as fallback. Triage retains a variant unless the evidence argues against its relevance and never discards variants in genes carrying two or more variants, in ACMG secondary-finding genes, or loss-of-function variants in phenotype genes. The model then compares the gene’s conditions with the phenotype and returns a verdict (YES, PARTIAL, INCIDENTAL, or EXCLUDED when the patient lacks a defining feature), which is passed unchanged to later stages (Figure 2). For each retained variant, the model writes a five-step reasoning and assigns ACMG/AMP criteria using a point-based rubric^33^: 10 points or more for Pathogenic, 6 to 9 for Likely Pathogenic, and 0 to 5 for uncertain significance. Variants of uncertain significance scoring 4 or 5 points are reported as variants of interest.

When parental genotypes are available, de novo, dominant, recessive, and X-linked layers are added. Segregation and phase are computed in code from allelic balance: values below 0.2 count as absent, 0.3 to 0.7 as heterozygous, and above 0.8 as homozygous, and a de novo call requires a heterozygous proband with both parents absent. Each layer copies the criteria of the previous one and adds only its own contribution: a de novo variant receives PS2 or PM6, and a variant inherited from an unaffected parent receives BS2. In recessive genes, a variant receives PM3 (+2) when its partner reaches the Likely Pathogenic threshold; the criterion is labeled a hypothesis when phase is unknown and an assumption when one allele is de novo. A homozygous proband receives PM3 at supporting strength (+1). For genes with two or more included variants, a cross-variant step evaluates whether they could act as a compound heterozygous pair.

#### Stage 4 (report)

Code assigns each variant to a report section (causative at 6 or more points or in a compound-heterozygous pair; variant of uncertain significance of interest at 4 to 5 points; ACMG secondary finding, v3.2, 81 genes, regardless of phenotype), and the model writes one text field per item through schema-constrained decoding. The report contains process details, augmented context, reasoning, and the final report.

#### Deterministic safeguards

Point totals are recomputed from the listed criteria after every layer; PMIDs and URLs absent from the retrieved evidence are removed; criteria whose eligibility depends on a tool verdict (PVS1, PS3, PM1, PM2, PM4, PP2/BP1, PP3, PS1/PM5, BP6, BS2) are added or stripped by validators; PP4 requires a full phenotype match and is stripped for genetically heterogeneous conditions; PS2 and PP1 come only from the inheritance layers; and a completeness check recovers any variant scoring at least 6 points that the summary omitted (Table S2).

### Evaluation

We test IVA with ten synthetic scenarios where trios present disease phenotypes including impaired vision, hypogonadotropic hypogonadism, congenital disorder of glycosylation, epilepsy, microchephaly, developmental delay and intellectual disability with different mode of inheritance. One intellectual disability trio carried a de novo variant in KDM2A, a gene recently discovered^34^ and not present yet in the genomic databases such OMIM or CGD. We then analyzed 16 trios solved by the Medical Genetics Service of Lausanne University Hospital (developmental delay, epilepsy, intellectual disability) and 327 solved singleton cases of visual impairment from the Jules-Gonin Eye Hospital, drawn from consanguineous families in Pakistan, for which the only phenotype provided was “visual impairment”. Ground truth was established by clinical geneticists blinded to the pipeline output. We assessed whether the expert-identified variant appeared in the final report and whether its ACMG class matched the expert classification. Sensitivity was the fraction of expert-reported variants classified as pathogenic, likely pathogenic, or of interest; precision, the fraction of pathogenic or likely pathogenic calls matching an expert-reported variant; and accuracy was computed over all candidate variants.

### Inference

In our system, Qwen3.5-9B ran in bfloat16 under vLLM (32,768-token context, 90% GPU memory, prefix caching) on one A100 (40 GB), with stage-specific thread pools of up to 64 workers for first-pass triage and 16 for GPU-bound reasoning stages. Model inference is local; evidence tools query public resources (NCBI, gnomAD, ClinGen, SpliceAI, AutoPVS1, UniProt, UCSC, CGD, Crossref) with single variant coordinates. A FastAPI service streams progress through server-sent events and answers clinicians’ follow-up questions with the same local model.

## Results

### Model selection

Among the local models, Qwen3.5-9B achieved the highest mean accuracy (0.812), strict accuracy (0.633), choice consistency (0.815), and inverted Brier score (0.875), whereas Qwen3.5-4B had the highest confidence consistency (0.958) (Table 1). Within the Qwen3.5 family, all metrics increased with the logarithm of model size (R^2^ above 0.75 for consistency and above 0.95 for accuracy and Brier score). Knowledge and robustness were correlated: no model combined high accuracy with low consistency, or the reverse (Figure 3). DeepSeek-V4-Pro, with 1.6 trillion parameters, outperformed every local model on every metric (mean accuracy 0.887, strict accuracy 0.713), but its margin over Qwen3.5-9B was modest given a roughly 180-fold difference in size. Qwen3.5-9B led on most topics except protein function and epigenetics, and the clinical MediPhi variants did not outperform general-purpose models of similar size (Figure S2). We therefore selected Qwen3.5-9B.

**Table 1.** Performance of 14 models on a 150-question clinical-genomics benchmark across five complementary metrics, evaluated under all 24 answer-option permutations per question. Confidence consistency and inverted Brier score are not reported for DeepSeek-V4-Pro, which does not expose token-level probability distributions; choice consistency is reported for all models. N Correct / Total indicates the number of questions on which the model was correct under all 24 permutations.

| Model | Size | Mean Acc | Strict Acc | Choice Cons. | Conf. Cons. | Brier Inv. | N Corr./Total |
| --- | --- | --- | --- | --- | --- | --- | --- |
| Gemma 4 E4B<sup>41</sup> | 8B | 0.729 | 0.533 | 0.757 | 0.901 | 0.797 | 80/150 |
| Llama 3.1 8B Instruct<sup>42</sup> | 8B | 0.650 | 0.433 | 0.653 | 0.913 | 0.771 | 65/150 |
| Phi-3 Mini 128k Instruct<sup>42</sup> | 3.8B | 0.639 | 0.413 | 0.672 | 0.921 | 0.755 | 62/150 |
| MediPhi-Clinical<sup>37</sup> | 3.8B | 0.584 | 0.267 | 0.568 | 0.710 | 0.648 | 40/150 |
| MediPhi-Guidelines<sup>37</sup> | 3.8B | 0.598 | 0.313 | 0.599 | 0.749 | 0.666 | 47/150 |
| MediPhi-Instruct<sup>37</sup> | 3.8B | 0.588 | 0.293 | 0.591 | 0.927 | 0.740 | 44/150 |
| Mistral 7B Instruct v0.3<sup>43</sup> | 7B | 0.487 | 0.153 | 0.315 | 0.371 | 0.510 | 23/150 |
| Qwen 2.5 3B<sup>44</sup> | 3B | 0.473 | 0.073 | 0.278 | 0.874 | 0.683 | 11/150 |
| Qwen 2.5 7B Instruct<sup>44</sup> | 7B | 0.722 | 0.467 | 0.746 | 0.815 | 0.753 | 70/150 |
| Qwen 3.5 0.8B<sup>45</sup> | 0.8B | 0.420 | 0.073 | 0.270 | 0.820 | 0.636 | 11/150 |
| Qwen 3.5 2B<sup>45</sup> | 2B | 0.610 | 0.313 | 0.620 | 0.927 | 0.752 | 47/150 |
| Qwen 3.5 4B<sup>45</sup> | 4B | 0.735 | 0.533 | 0.764 | 0.958 | 0.824 | 80/150 |
| **Qwen 3.5 9B<sup>45</sup>** | **9B** | **0.812** | **0.633** | **0.815** | **0.957** | **0.875** | **95/150** |
| DeepSeek-V4-Pro<sup>46</sup> | 1.6T | 0.887 | 0.713 | 0.880 | NR | NR | 107/150 |

**Figure 3.**
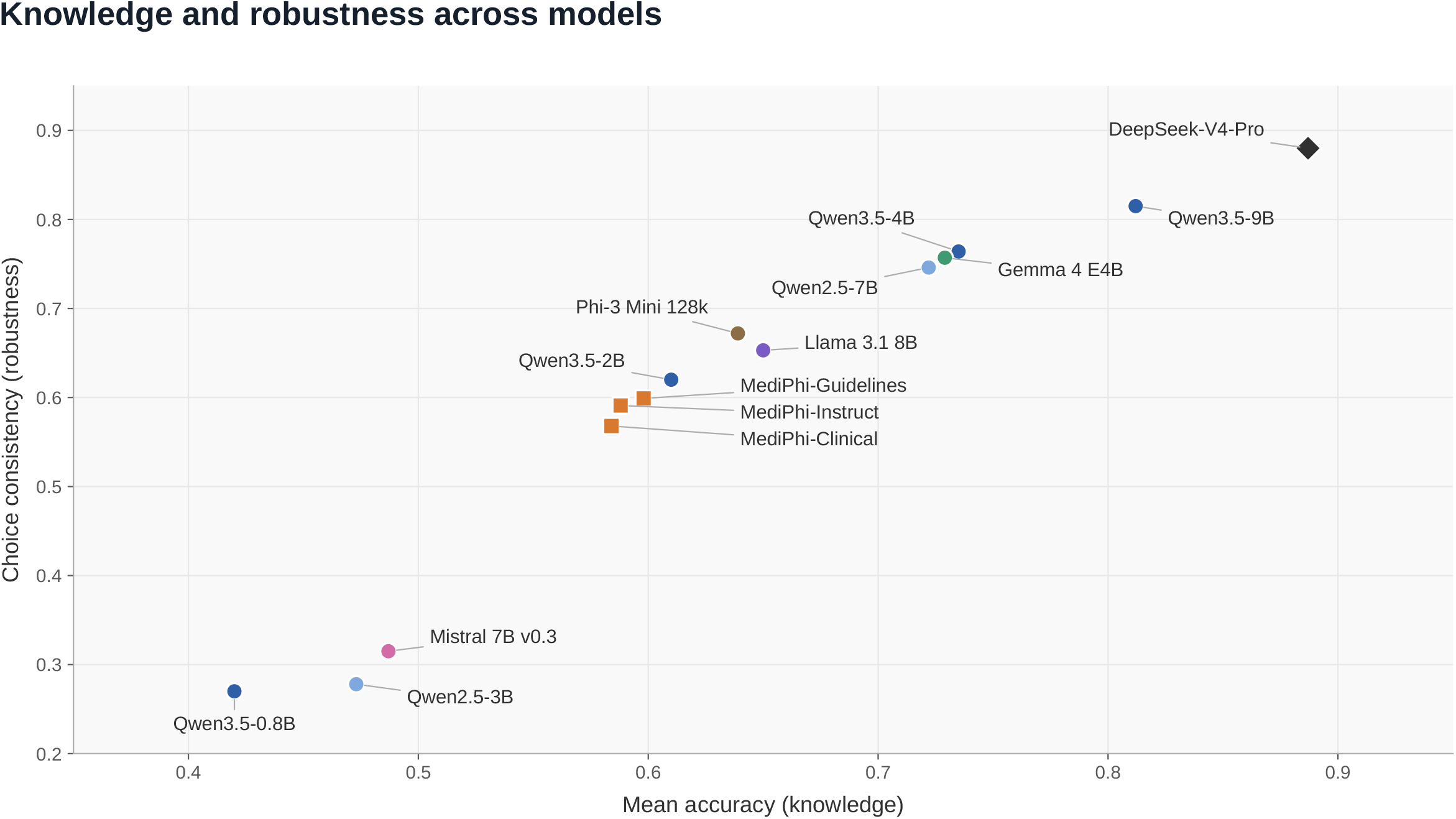
Knowledge and robustness across models. Mean accuracy (knowledge, x-axis) against choice consistency (robustness, y-axis) for the 13 locally evaluated SLMs and DeepSeek-V4-Pro. Models lie along a diagonal envelope; none combines high knowledge with low robustness or the reverse.

### Synthetic scenarios

Across the synthetic trios (6–7 variants each), IVA reported the expected causative variant in all the cases, each as Pathogenic or Likely Pathogenic. De novo variants in CHD2, EBF3, KDM2A and MECP2 were classified Pathogenic (11–16 points). The homozygous WWOX splice-site variant (both parents carriers) and the hemizygous RS1 variant inherited from a carrier mother were classified Pathogenic (12 and 15 points). In the KDM2A trio (6 variants, both parents negative), IVA retrieved the gene–disease link from recent literature, applied PVS1 (+8), PS2 (+4), PM2 (+2) and PP4 (+1), and classified the nonsense variant as Pathogenic (15 points). In the same trio it discarded an unrelated and neutral MEN1 variant and excluded a single heterozygous variant in the recessive gene RPE65. A TP53 missense variant inherited from an unaffected father was downgraded by BS2 to uncertain significance (4 points), and an SCN5A variant was reported as a secondary finding. In hypogonadotropic hypogonadism, IVA set aside phenotype-incongruent variants (RHO, BRCA1, MYBPC3, FLNA, VWF) and identify two variants in GNRHR. Because the two GNRHR variants came from different parents, it treated them as compound heterozygous in trans (PM3 +2) and classified the nonsense allele as Pathogenic (10 points) and the missense allele as Likely Pathogenic (7 points). PMM2 was handled the same way when both alleles were inherited (11 and 17 points). In the trio where one PMM2 allele was set as de novo, IVA assumed the trans configuration and stated that phase must be checked.

### Real diagnostic cohorts

Of 16 solved trios, eleven were explained by de novo variants (SMAD4, ARID1B, SRCAP, MBD5, CDK13, CACNA1C, SMARCA2, and TUBA4A among them, with a de novo variant of uncertain significance in GRIN2A in one case), four by inherited variants (TCF7L2 and X-linked SYN1 from a mosaic mother, FGFR3 paternal, SRRM2 maternal), and one by compound-heterozygous RYR1; a maternal BRCA1 variant was reported as a secondary finding. IVA reported every previously identified pathogenic or likely pathogenic variant with the expert ACMG class (sensitivity 100%), with 2.8±1.4 P/LP calls per case (precision 39.5%; accuracy 98.2% over all candidates). Excess calls arose in two situations: likely artifactual variants with a low proband allelic ratio called de novo (we are supposing QC filtering happening upstream), and variants inherited from a presumably unaffected parent for which strong molecular evidence outweighed the BS2 penalty and the model invoked reduced penetrance without a reference.

In 327 singleton cases, IVA surfaced 93% of the expert-reported variants, with substantial agreement in ACMG scoring: 82% as pathogenic or likely pathogenic and 11% as variants of uncertain significance of interest. For pathogenic or likely pathogenic calls, sensitivity was 82%, precision 43%, and accuracy 99%, with 1.9±1.2 calls per case (Table 2). Remaining cases (7%) were retrieval misses: in almost all missed cases the causative gene was identified during the analyses but not surfaced in the report, and in one the cause was not recovered.

**Table 2.**
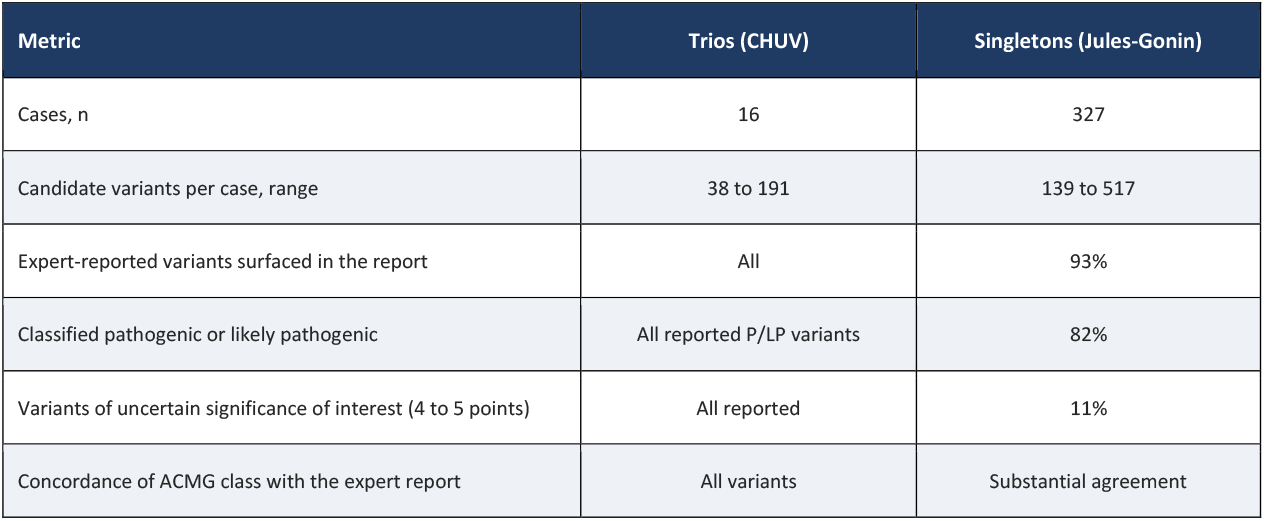
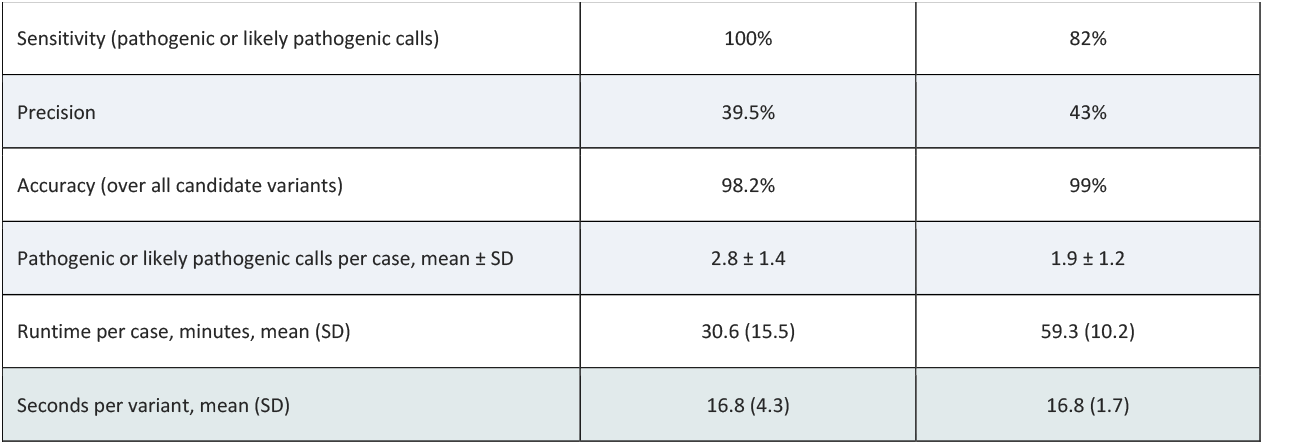
Performance in two real diagnostic cohorts. Sensitivity, precision, and accuracy refer to pathogenic or likely pathogenic calls compared with the variants reported by the expert analysts (Methods).

| Metric | Trios (CHUV) | Singletons (Jules-Gonin) |
| --- | --- | --- |
| Cases, n | 16 | 327 |
| Candidate variants per case, range | 38 to 191 | 139 to 517 |
| Expert-reported variants surfaced in the report | All | 93% |
| Classified pathogenic or likely pathogenic | All reported P/LP variants | 82% |
| Variants of uncertain significance of interest (4 to 5 points) | All reported | 11% |
| Concordance of ACMG class with the expert report | All variants | Substantial agreement |
| Sensitivity (pathogenic or likely pathogenic calls) | 100% | 82% |
| Precision | 39.5% | 43% |
| Accuracy (over all candidate variants) | 98.2% | 99% |
| Pathogenic or likely pathogenic calls per case, mean ± SD | 2.8 ± 1.4 | 1.9 ± 1.2 |
| Runtime per case, minutes, mean (SD) | 30.6 (15.5) | 59.3 (10.2) |
| Seconds per variant, mean (SD) | 16.8 (4.3) | 16.8 (1.7) |

### Failure analysis and throughput

Testing on synthetic cases revealed four recurrent failure classes, each addressed by a safeguard (Supplementary Appendix): i) omission of a correctly scored variant from the summary; ii) criteria granted without verifying the mechanism; iii) correct disqualifying reasoning followed by an uncapped pathogenic label; and iv) arithmetic or notation drift introduced during compression. Runtime averaged 59.3 minutes (SD 10.2) per singleton case (139 to 517 variants; r = 0.80 with case size) and 30.6 minutes (SD 15.5) per trio (38 to 191 variants), about 16.8 seconds per variant in both sets, on one GPU.

### Discussion

This study shows that a small language model of 9 billion parameters, running on a single GPU within the hospital network, can generate complete, ACMG-aligned reports for real diagnostic cases. Among 139 to 517 candidate variants per case, IVA recovered nearly all causative variants previously identified by expert analysts, at a throughput compatible with routine laboratory work. In existing systems, language models retrieve and summarize evidence while a deterministic engine performs the classification. In IVA, the model carries out the interpretive reasoning itself: it assigns ACMG criteria, integrates the evidence with zygosity and parental genotypes, and draws a case-level conclusion. In trios, it applied de novo, dominant, recessive, and X-linked inheritance models, and when phase could not be determined, it reported compound heterozygosity as a hypothesis requiring confirmation rather than as an established finding.

This performance depends on the synergy between the deterministic harness built around the model and the prompting structure. Small models are unreliable at arithmetic, verbatim copying, and strict formatting, so these tasks were removed from the model wherever possible. The model proposes criteria and their strengths, but totals, layer-by-layer sums, and classes are recomputed in code. Any PMID or URL absent from the retrieved evidence is removed, and each text field of the final report can cite only identifiers that the code supplied for it. The base criteria are transferred into each inheritance layer by the code rather than retyped by the model, and criteria that depend on a tool result are added or removed by validators that compare the model’s choices with the tool output. The format of the report is enforced at three levels: the code renders the section headers and criterion lines, several calls use schema-constrained decoding, and if a model call fails, the report is generated from the retrieved facts alone. Each safeguard answered a specific failure, and some initially failed silently, for example when a regular expression did not match the layout the model actually produced.

Despite its framework, IVA is not deterministic. All calls use greedy decoding, yet continuous batching, concurrent requests, reduced-precision arithmetic, and prefix caching make token-level output depend on batch composition. Evidence tools query live public resources that are updated over time or occasionally rate-limit or time out. Repeated runs of the same case can therefore differ in wording, in the retrieved evidence, and occasionally in a secondary finding or a borderline classification.

Several limitations apply. Evaluation was retrospective, at two centers in one city, and the outcome measures were recovery of expert-reported variants and agreement with expert classes, not the quality or usefulness of the reports, nor a comparison with analyst time or accuracy. Precision was low (39.5% and 43%) because we favored sensitivity; analysts must still review every call. The singleton cohort comes from consanguineous families enriched for homozygous recessive disease. The benchmark ranks models but is not a validated instrument, and multiple-choice performance is an imperfect proxy for clinical clinical competence^35,36^. Adversarial or malformed inputs have not been tested, and parental allelic balance is used without read depth, so a poorly covered parent can mimic a de novo variant; we retain such calls and recommend Sanger confirmation. Finally, the evidence tools send variant coordinates to public servers, a dependency that local database snapshots would remove.

Future work includes external validation, participation in quality assessment schemes such as EMQN, comparison with analysts, and distillation or quantization for less powerful hardware. Models with domain-specific pretraining are another option, although the specialized MediPhi versions^37^ did not outperform general-purpose SLMs and Meditron3^38^ has not yet been tested. The manifest-based architecture of IVA can be extended to copy-number variants^39^ and long-read structural variants^40^, and to richer evidence resources as provided by the MARRVEL-MCP interface.

In conclusion, a small language model running on local hardware can serve as the reasoning layer for case-level and trio-based variant interpretation, provided it is surrounded by deterministic safeguards. IVA remains a decision-support tool: its reports require review by an analyst, and its performance must be confirmed in prospective studies.

## Data Availability

The IVA source code, tool manifests, prompts, and client-server interface are available at https://github.com/fredsanto/IVA under the GNU Affero General Public License v3.0. The 150-question benchmark and its permutation scoring script are provided in the same repository; the synthetic genetic profiles contain no true patient data. Patient-derived profiles from the CHUV and Jules-Gonin cohorts cannot be shared because they contain identifiable genetic information and fall outside the scope of the original 

## Funding

This work has been supported by the EU Horizon 2020 Research and innovation Program N. 847941 (miniNO) to F.S. and by the Swiss National Foundation (grant N.10.009.066) to F.S. and M.A. and by the Guillaume-Gentil Foundation to C.L.C.

## Competing interests

The authors declare no competing interest.

## Ethics

The use of patient data in this study was approved by the ethics committees of participating institutions in Pakistan (F.No.HU/ORIC/2026/160; No.3837/ORIC) and Switzerland (CER-VD 2021-01400).

## Data and code availability

The IVA source code, tool manifests, prompts, and client-server interface are available at https://github.com/fredsanto/IVA under the GNU Affero General Public License v3.0. The 150-question benchmark and its permutation scoring script are provided in the same repository; the synthetic genetic profiles contain no true patient data. Patient-derived profiles from the CHUV and Jules-Gonin cohorts cannot be shared because they contain identifiable genetic information and fall outside the scope of the original consent.

## Author contributions

F.S. conceived the study, developed the system, analyzed the data, wrote the manuscript, and supervised the project. E.D. developed the benchmark questionnaire, analyzed the data, developed the system, and wrote the manuscript. J.Z. contributed to development of the system. S.O. contributed to clinical sample retrieval and processing. A.R., M.U., H.F., and M.A. provided the visual-impairment cohort data., A.L., R.F., and C.L.C. provided the trio cohort data. All authors reviewed and approved the final manuscript.

## Acknowledgments

We thank Prof. Alexandre Reymond (UNIL) for his observations and suggestions, and Florence Niel-Bütschi (CHUV) for supervising the clinical case selection from the Medical Genetics Service of the CHUV. Artificial intelligence (Claude, a large language model) was used to assist with coding and language editing (spelling and syntax); the authors reviewed and take responsibility for all content.

## Notes

### Competing Interest Statement

The authors have declared no competing interest.

### Author Declarations

The use of patient data in this study was approved by the ethics committees of participating institutions in Pakistan (F.No.HU/ORIC/2026/160; No.3837/ORIC) and Switzerland (CER-VD 2021-01400). 1. HU/ORIC/2026/160: IRB of Hazara University Mansehra, KPK, Pakistan 2. No.3837/ORIC: Institutional Bioethics Committee (IBC) of the Islamia University of Bahawalpur, Pakistan 3. CER-VD 2021-01400: Commission cantonale (VD) d’éthique de la recherche sur l’être humain

