## Supplementary material for "IVA, a locally deployable Intelligent Variant Analyser based on Small Language Model for complete case-level and trio-based genomic interpretation": Table S1

This appendix contains extended methods, the deterministic validators, the failure analysis, and supplementary figure legends referred to in the main text. It contains no reference list; citations will be added through EndNote.

#### 1. Supplementary Methods

##### 1.1 Input, normalization, and run settings

The input is a table in CSV, TSV (delimiter detected automatically) or Excel format and a free-text phenotype. A model call interprets the column headers and maps them to a canonical schema (variant identifier, coordinates, reference and alternate alleles, type, transcript, HGVS, zygosity, gene, OMIM phenotype and inheritance, ClinVar class, allelic balance of proband (mandatory), mother and father (if available), frequency, and in-silico scores such as CADD, REVEL, SIFT, PolyPhen-2, AlphaMissense and SpliceAI); a human-readable summary of the mapping is returned to the user. The genome build is detected from the variants. All model calls use temperature 0 with extended reasoning disabled, because reasoning tokens interfered with downstream parsing. Maximum output tokens are set per stage.

##### 1.2 Stage 1 evidence tools

**Table S1. ACMG Evidence Tools**

| Tool | Source | Role and gating |
| --- | --- | --- |
| SpliceAI | Public API (hg19 and hg38 endpoints), MANE Select transcript | Delta scores; skipped when splicing is irrelevant or the input already has a SpliceAI column |
| gnomAD frequency and constraint | gnomAD API (v4 for hg38; v2.1.1 for hg19) | Population frequency (PM2, BA1/BS1 context), pLI and LOEUF |
| ClinVar gene statistics, residue search, hotspot | NCBI E-utilities | Gene-level missense versus truncating verdict (PP2/BP1), same-residue and same-change hits (PS1/PM5), hotspot window (PM1), submission-level tags for functional or case evidence (PS3, case reference) |
| ClinGen Allele Registry | reg.clinicalgenome.org | Allele registration and cross-references |

| | | |
|---|---|---|
| GeneReviews, MedGen features, gene function | NCBI | Gene-level condition lists, clinical features of each condition, gene function |
| UniProt domain, repeat region | UniProt REST, UCSC API | Domain evidence for PM1; repeat check for PM4 |
| AutoPVS1 | Public AutoPVS1 server, behind a hybrid gate | PVS1 strength and decision path, pLI and haploinsufficiency; an ambiguous gate is resolved by a one-token model call |
| LitVar2 and PubMed | NCBI | Variant-level track tied to the phenotype; patient-blind gene-level track based on OMIM or CGD conditions; functional-evidence and case-reference tags with PMIDs |
| Literature agent | PubMed (E-utilities) and Crossref (bioRxiv, medRxiv) | Runs only when GeneReviews, OMIM phenotype, MedGen and CGD are all empty for the gene; titles scored by the model, top abstracts summarized; the patient phenotype is never used |

Each tool output is optionally compressed (threshold of ten variants by default and twenty for AutoPVS1, LitVar2, SpliceAI, and the literature agent). The compressor must keep section headers, ACMG codes with strengths, HGVS notation, classifications, gene-disease statements including negative findings, and citation URLs; on failure a character-level truncation with an explicit marker is used.

#### 1.3 Triage, phenotype verdict, and reasoning

First triage returns a JSON object with the strongest argument to keep, the strongest argument to discard, and a decision. A documented gene-disease link, variant-specific LitVar2 evidence, VUS, missing or conflicting evidence argue for keeping; an explicit absence of disease link, or a ClinVar Benign or Likely Benign classification without conflict, argue for discarding. If the evidence links the gene to at least one distinct cluster of the phenotype, the variant is kept. Overrides prevent discard of genes with two or more variants (compound-heterozygous candidates), ACMG secondary-finding genes, and loss-of-function variants in phenotype genes.

A patient-blind call lists every condition attributed to the gene. A second, patient-aware call compares that list with the phenotype and returns, for each condition, MATCH, NO MATCH or EXCLUDED (a core defining feature is explicitly absent in the patient), and a verdict: YES (a matching condition covers the phenotype as a whole), PARTIAL, INCIDENTAL (no match and no excluded condition), or EXCLUDED. The list and verdict are stored per variant and reinjected unchanged into later stages. Retained variants receive a five-step reasoning (phenotype fit with citations, notation, molecular mechanism, inheritance coherence, ambiguities) that forbids uncited external claims and premature classification.

#### 1.4 Deterministic validators of ACMG/AMP criteria

The model proposes criteria with a strength and points (Supporting 1, Moderate 2, Strong 4, Very Strong 8; benign criteria negative; BA1 stand-alone -8). A recognizer reads criterion codes and strengths from the free text, ignores negated or prose mentions, and feeds the totals. Classification thresholds: Pathogenic 10 or more, Likely Pathogenic 6 to 9, uncertain significance 0 to 5, Likely Benign -6 to -1, Benign -7 or less.

**Table S2. Deterministic validators of ACMG/AMP criteria and totals.**

| Criterion | Rule enforced in code |
|---|---|
| PVS1 | Stripped when AutoPVS1 returns 'not applicable'; an unresolved AutoPVS1 verdict cannot yield Very Strong PVS1 from gene constraint alone; gated on phenotype fit |
| PS3 | Allowed only from tool-side tags: a ClinVar P/LP submitter comment reporting functional work with a PMID, or variant-level functional evidence from LitVar2 with a retrieved PMID; the PMIDs are carried into the final report |

| | |
|---|---|
| PS1 / PM5 | Added when the residue search shows the same (PS1) or a different (PM5) amino-acid change classed Likely Pathogenic or above at the residue and the model omitted it |
| PS2 / PP1 | Excluded from the base score; contributed only by the inheritance layers |
| PM1 | Kept or added only when the ClinVar hotspot window or the UniProt domain rule is met; otherwise stripped |
| PM2 | Added when the allele frequency is below 1e-4 or the variant is absent from gnomAD; never stripped (a strip rule would have removed PM2 from 50 variants of the 327-report corpus, including 40 in recessive genes) |
| PM3 | Set by code in the recessive layers (see 1.5) |
| PM4 | Added for in-frame indels outside repeat regions and stop-loss variants; stripped otherwise and never combined with PVS1 |
| PP2 / BP1 | Kept only for missense variants when the gene-level ClinVar verdict (missense-predominant or truncating-predominant) matches exactly |
| PP3 | Kept only when at least two in-silico tools cross their deleterious thresholds (REVEL 0.7, CADD 20, SIFT 0.05, PolyPhen-2, AlphaMissense, SpliceAI 0.5); SpliceAI alone suffices for splice-impact variants |
| PP4 | Kept only when the phenotype verdict is YES; stripped when the justification names a condition on a maintained list of genetically heterogeneous disorders (for example familial hypercholesterolemia) |
| BP6 | Stripped when the model's own justification states that the ClinVar class does not meet the criterion |
| BS2 | Applied by default to a variant inherited from an unaffected parent in dominant genes, and when a parent is homozygous in the recessive homozygous path, unless the transmitting parent is stated to be affected |
| ClinVar case reference | One Supporting criterion (+1; PM3 or PS4_Supporting) added when a P/LP submission cites a PMID that is case rather than functional evidence |
| Totals | Re-summed from the listed criteria after every layer; the stated total and class are overwritten when they disagree; base criteria are copied by code into each layer |

#### 1.5 Segregation, phasing, and recessive criteria

Segregation is computed from allelic balance (AB). A value below 0.2 is absent, 0.3 to 0.7 heterozygous, above 0.8 homozygous, and values in the gaps are uncertain. De novo requires a heterozygous proband and both parents absent (proband AB at least 0.35 when a parent shows a trace read of 0.02 to 0.2, which also triggers a possible mosaicism note). Other outcomes are maternal, paternal, both carriers, homozygous parent, uncertain, or insufficient data; each value is defined explicitly in the prompts because the model once misread the homozygous-parent value as supporting a de novo call. Two variants in a gene are cis when inherited from the same parent, trans when from different parents, and unknown otherwise; if one is de novo, trans is assumed and the report must state that phase has to be checked.

PM3 is set by code. For a compound-heterozygous pair, a variant receives PM3 (+2) when its partner reaches at least 6 points on its own base score: firm under confirmed trans, labeled as assumed when one allele is de novo, and labeled a hypothesis (with the class prefixed Potential) when phase is unknown. A partner below 6 points gives +0. For a homozygous proband PM3 is applied at supporting strength (+1; ClinGen SVI counts one homozygous proband as 0.5 case points), with no reduction for presumed consanguinity, and is withheld when a parent is homozygous.

#### 1.6 Secondary findings

Every variant is checked against the ACMG SF v3.2 list (81 genes). A P/LP variant in one of these genes is protected from discard regardless of phenotype or of which family member carries it, labeled INCIDENTAL when the gene is unrelated to the phenotype, and reported in a dedicated section that states the gene, the condition and that the finding is reported independently of the primary phenotype.

#### 1.7 Report construction

The output has four sections: process details, augmented context, reasoning, and final report. In the final report, code decides the section of each variant: causative (phenotype verdict YES or PARTIAL, included by the second triage on its own, and a total of at least 6 points or a causative compound-heterozygous pair), variant of uncertain significance of interest (4 to 5 points), or secondary finding. The model writes one text field per item and returns it as JSON under a schema, so it cannot add, drop or move an item; code removes cited identifiers (PMID, OMIM, GeneReviews, MedGen, UniProt, ClinVar and SCV identifiers) that it did not supply for that item and sentences that name another gene of the run, copies the criteria lines verbatim, and renders fixed headers. If the model call fails, the report is rendered from the facts alone. A deterministic completeness check appends any variant meeting the causative threshold that the conclusion omitted, and a focused model pass then tries to integrate it.

#### 1.8 Inference and interactive layer

Qwen3.5-9B (bfloat16) is served by vLLM with continuous batching, a 32,768-token context, 90% GPU memory utilization and prefix caching on one A100 (40 GB). A FastAPI service exposes endpoints to submit an analysis (/analyze), stream progress as server-sent events (/stream/{job}), cancel, retrieve the result, and chat with the report (/chat/{job}); the chat is answered by the same local model.

#### 1.9 Benchmark metrics

Confidence consistency is one minus the mean variance of an option's probability across the 24 orderings, normalized by its theoretical maximum of 3/16, the variance of a Bernoulli variable with probability 1/4. Choice consistency applies the same formula to the discrete answer. The inverted Brier score is 1 - BS/2. At temperature 0 a cloud model returns a single answer instead of a probability vector, so its inverted Brier score reduces to mean accuracy and is not reported. The stored answer key of the 150 questions is not balanced (B 60%, C 21%, A 16%, D 3%), which the permutation design neutralizes.

### Figure S1

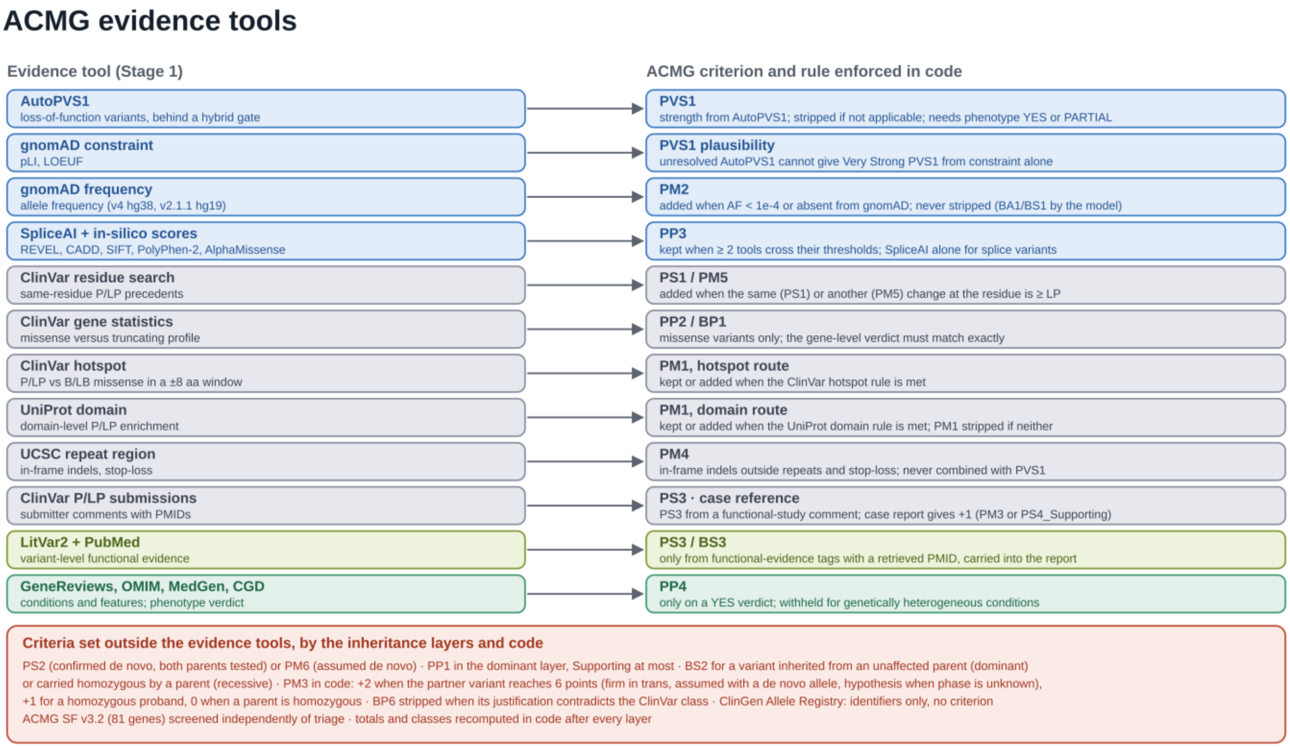


**Figure S1.** Each Stage 1 evidence tool (left) is linked to the ACMG/AMP criterion it grounds and to the rule that code enforces on the model's choice (right). Colors match the tool groups in Figure 1. The box at the bottom lists criteria assigned by the inheritance layers or by code without a dedicated tool. In addition the model can propose BA1, BS1 and BP4. AF denotes allele frequency; LP, likely pathogenic; P/LP, pathogenic or likely pathogenic; and B/LB, benign or likely benign. Details are given in Table S1.

#### Figure S2

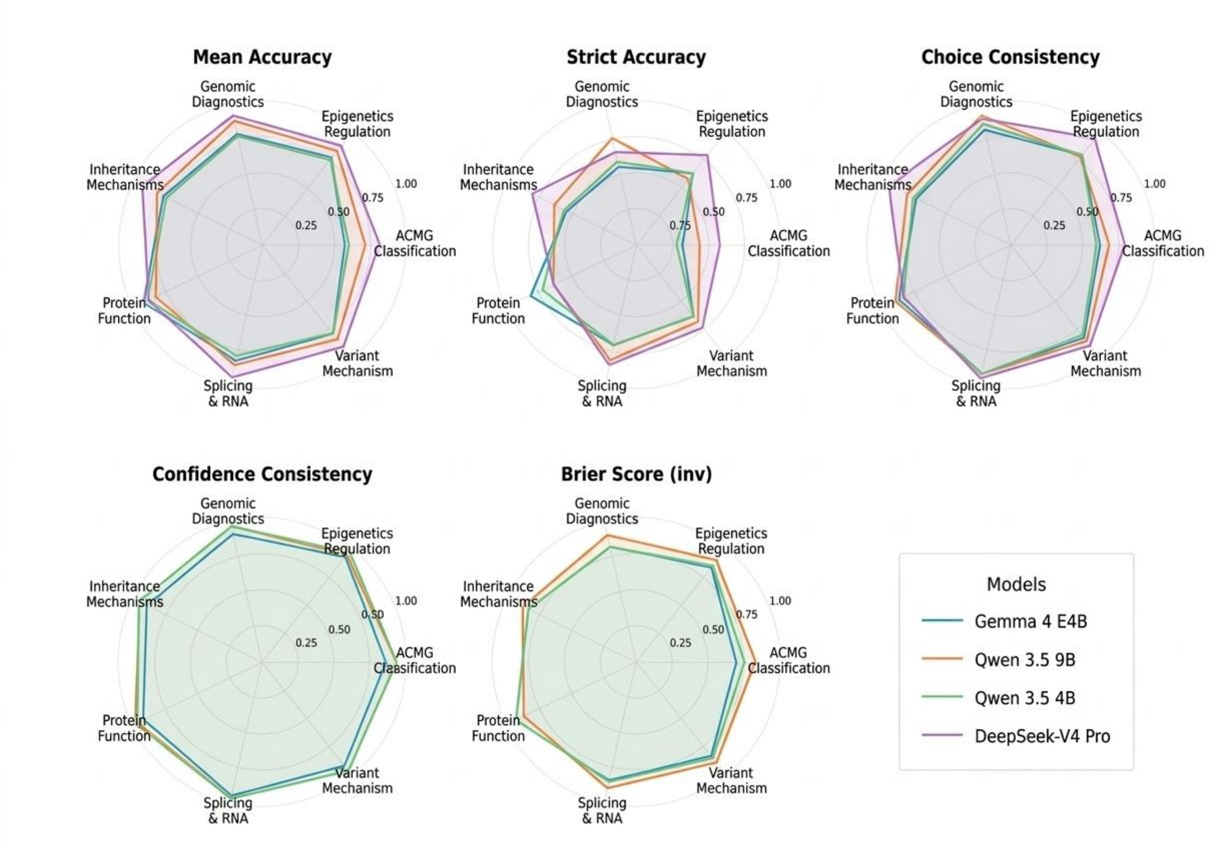


**Figure S2.** Per-topic breakdown of all five evaluation metrics for the four best-performing models, displayed as radar charts. Each axis represents a clinical-knowledge competence label; radial distance indicates metric value (0 to 1). Confidence consistency and inverted Brier score are omitted for DeepSeek-V4-Pro, which does not expose token-level probability distributions.
